# Tracking the progressive spread of the SARS-CoV-2 Omicron variant in Italy, December 2021 - January 2022

**DOI:** 10.1101/2022.01.27.22269949

**Authors:** Paola Stefanelli, Filippo Trentini, Daniele Petrone, Alessia Mammone, Luigina Ambrosio, Mattia Manica, Giorgio Guzzetta, Valeria d’Andrea, Valentina Marziano, Agnese Zardini, Carla Molina Grane, Marco Ajelli, Angela Di Martino, Flavia Riccardo, Antonino Bella, Monica Sane Schepisi, Francesco Maraglino, Piero Poletti, Anna Teresa Palamara, Silvio Brusaferro, Giovanni Rezza, Patrizio Pezzotti, Stefano Merler, the Genomic SARS-CoV-2 National Surveillance Working Group, the Italian Integrated Surveillance of COVID-19 Study Group

## Abstract

The SARS-CoV-2 variant of concern Omicron was first detected in Italy in November 2021. Data from three genomic surveys conducted in Italy between December 2021 and January 2022 suggest that Omicron became dominant in less than one month (prevalence on January 3: 78.6%-83.8%) with a doubling time of 2.7–3.1 days. The mean net reproduction number rose from about 1.15 in absence of Omicron to a peak of 1.83 for symptomatic cases and 1.33 for hospitalized cases, while it remained stable for critical cases.

## Main text

The SARS-CoV-2 variant of concern (VOC) Omicron, characterized by a large number of mutations, has shown a marked growth advantage over pre-circulating lineages, causing a major upsurge of cases in multiple countries since November 2021 [1]. As Europe transitions rapidly from Delta to Omicron dominance, data on presence and prevalence of VOCs across distinct geographic contexts and over time can support assessments of the current and potential short-term implications for public health. Here, we report the results of three quick genomic prevalence surveys conducted biweekly in Italy between December 6, 2021, and January 3, 2022. The estimated prevalence of Omicron is combined with data gathered by the Italian Integrated Surveillance System to assess the impact of the variant spread on the overall circulation of SARS-CoV-2.

### The spread of the Omicron variant in Italy

Genomic prevalence surveys were conducted on December 6 and 20, 2021, and January 3, 2022. The initiative involved all 19 Regions and 2 Autonomous Provinces (APs) of Italy and was coordinated by the National Public Health Institute (Istituto Superiore di Sanità, ISS), in collaboration with the Ministry of Health, the Bruno Kessler Foundation, and 120 laboratories distributed across the national territory. Random samples were selected in each Region/AP and uniformly distributed across 4 macro-areas (North-East, North-West, Center, and South/Islands) among infections diagnosed with a real-time reverse transcription polymerase chain reaction (RT-PCR) on the dates of the surveys. The sample size was calculated to enable the detection of circulating SARS-CoV-2 variants with at least 5% prevalence within each macro-area with a 2% precision. Samples were subjected to whole genome sequencing (WGS) as the gold standard method for variants detection. Alternatively, results from Sanger or next generation sequencing (NGS) of the whole or partial S-gene were collected. Sequences with insufficient quality for variant assignment were discarded from the analysis. To robustly assess the temporal expansion of Omicron, the prevalence of this variant was estimated in each Region/APs using two alternative approaches. The first used Markov Chain Monte Carlo (MCMC) under the assumption of independence in prevalence across Regions/APs and surveys. The second fitted a generalized linear mixed model (GLMM) assuming a random intercept and a random slope for each Region/APs; the independent variable was the day when the survey took place. A binomial distribution for the identified number of Omicron sequences was assumed in both approaches. The procedures were replicated on data aggregated by macro-area.

On December 6, 2021, 2241 samples were collected and 2127 successfully sequenced; of these 4 resulted Omicron (0.2% of samples) and 2121 Delta (99.7%). Omicron was only found in Northern Italy (Emilia-Romagna, Lombardy and Veneto). Estimates of the national prevalence ranged from 0.2% to 2.1%.

On December 20, 2021, 2194 samples were collected and 2139 successfully sequenced; 462 resulted Omicron (21.6%) and 1676 Delta (78.4%). Omicron was detected in all Regions/APs, and it was the predominant variant in Umbria (Central Italy). Estimates of the national prevalence ranged from 17.1% to 23.5%.

By January 3, 2022 (2632 samples, 2571 successfully sequenced), Omicron had become largely dominant in the country, being confirmed in 2058 (80.0%) sequences (Delta: 512 sequences, 19.9%). Estimates of the national prevalence ranged from 78.6% to 83.8%. Results for the three surveys are reported in Figure 1 and Table 1. Corresponding prevalence estimates are reported in Table 2.

**Table 1.**
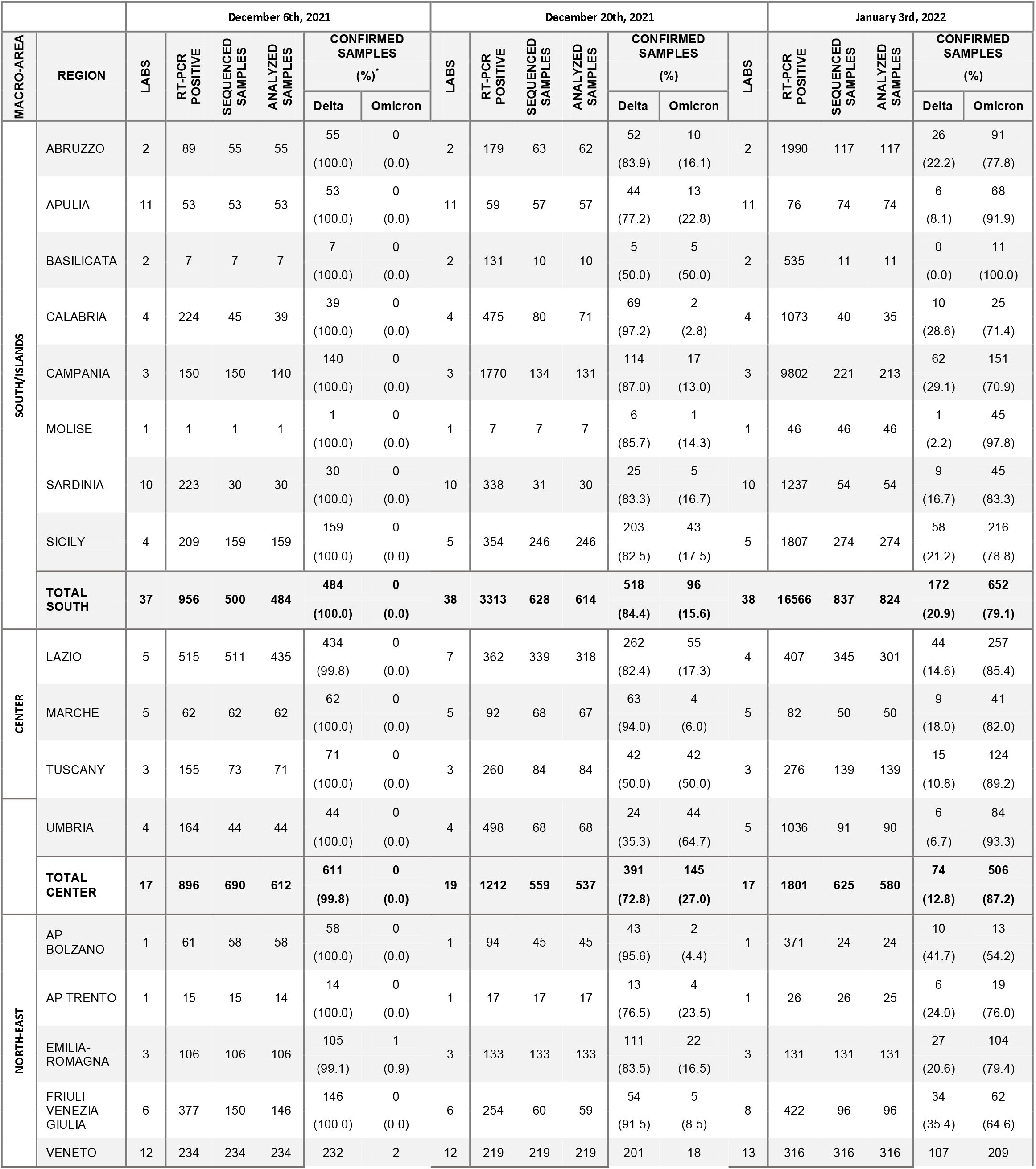
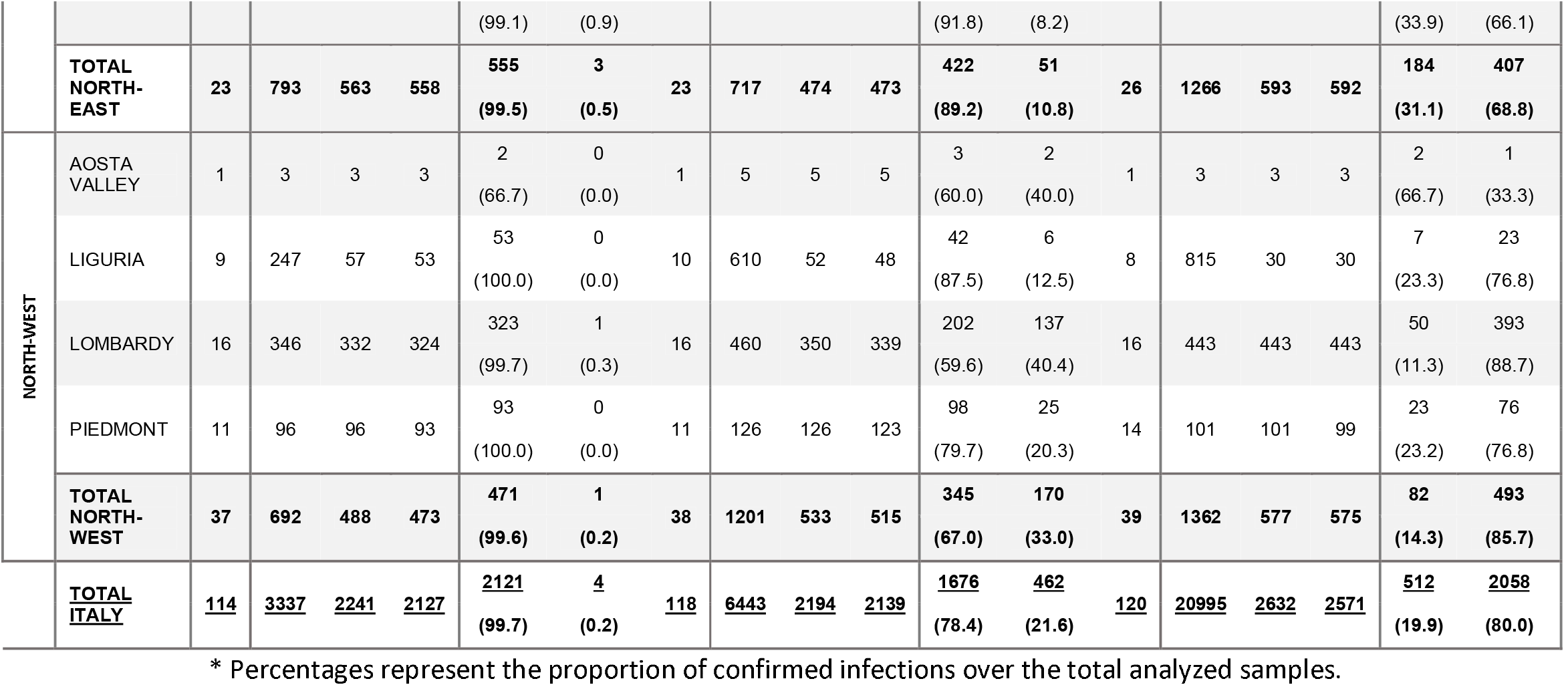
Results across the 21 participating Regions/APs.

**Table 2.** National level estimates from surveys data for the Omicron prevalence, exponential growth rate, doubling time and net reproduction number.

|  |  | GLMM |  | MCMC |  |
| --- | --- | --- | --- | --- | --- |
|  |  | By Region/AP | By macro area | By Region/AP | By macro area |
| prevalence (%) | December 6th, 2021 | 1 (0.8 - 1.4) | 1.1 (0.8 - 1.4) | 1.4 (0.9 - 2.1) | 0.5 (0.2 - 0.9) |
|  | December 20th, 2021 | 18.7 (17.1 - 20.4) | 19.1 (17.5 - 20.8) | 21.5 (19.7 - 23.5) | 21.5 (19.7 - 23.3) |
|  | January 3rd, 2022 | 82.2 (80.6 - 83.8) | 81.3 (79.7 - 82.8) | 80.3 (78.6 - 81.9) | 80.1 (78.6 - 81.6) |
| r (days<sup>-1</sup>) |  | 0.228 (0.173 - 0.285) | 0.225 (0.171 - 0.282) | 0.216 (0.162 - 0.276) | 0.26 (0.183 - 0.346) |
| T (days) |  | 3.1 (2.44 - 4) | 3.13 (2.46 - 4.05) | 3.27 (2.51 - 4.29) | 2.73 (2.01 - 3.79) |
| R if GT=4 days |  | 1.91 (1.69 - 2.14) | 1.9 (1.68 - 2.13) | 1.86 (1.65 - 2.1) | 2.04 (1.73 - 2.38) |
| R if GT=6 days |  | 2.37 (2.04 - 2.71) | 2.35 (2.03 - 2.69) | 2.3 (1.97 - 2.65) | 2.56 (2.1 - 3.07) |
| R if GT=8 days |  | 2.82 (2.39 - 3.28) | 2.8 (2.37 - 3.26) | 2.73 (2.29 - 3.21) | 3.08 (2.46 - 3.76) |

**Figure 1.**
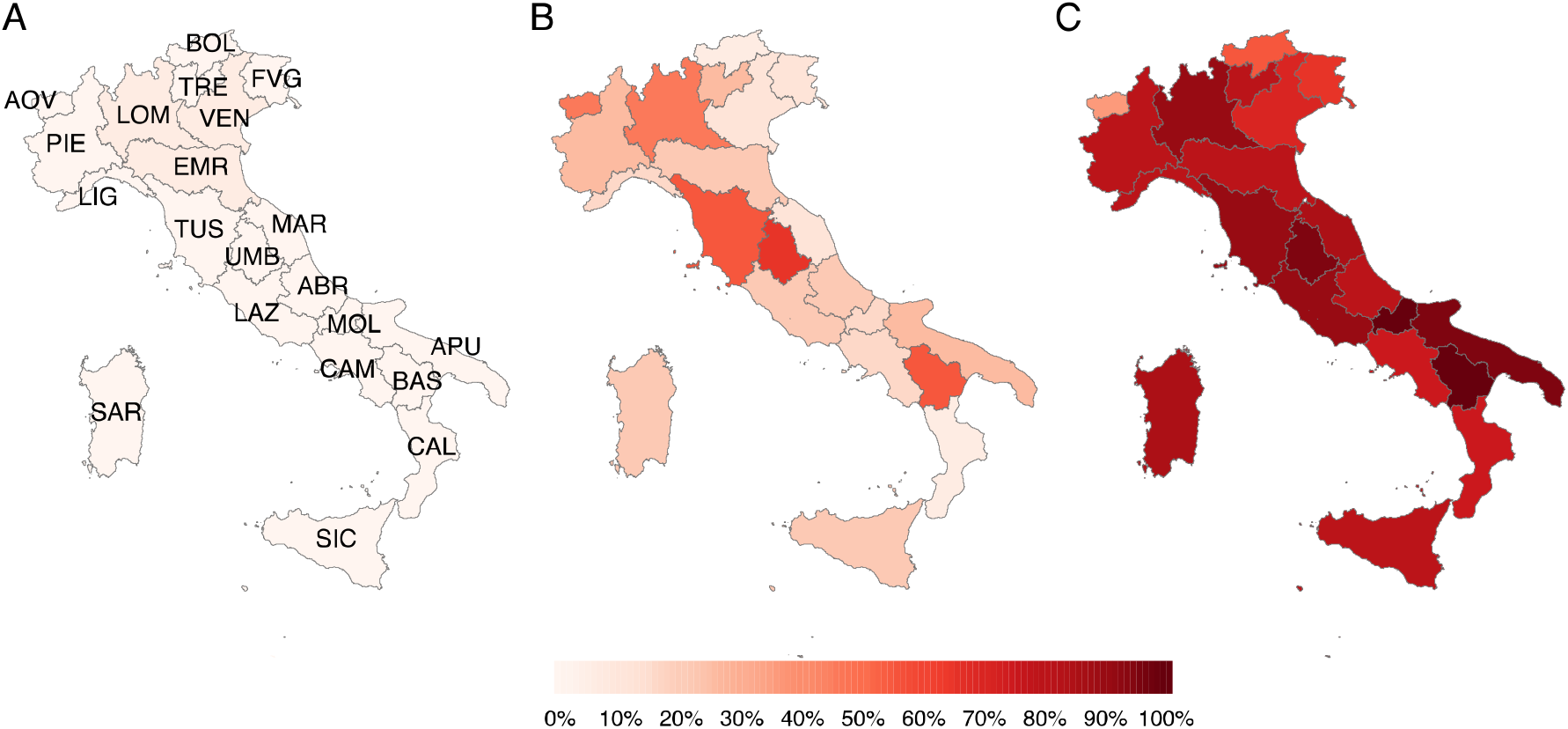
Geographical distribution of SARS-CoV-2 variant Omicron. Point prevalence of Omicron by the 19 Regions and 2 Autonomous Provinces of Italy as obtained from national surveys conducted on December 6, 2021, December 20, 2021, and January 3, 2022. Abbreviations of region names are reported in the leftmost panel and listed in the Appendix.

Multiplying the prevalence in each Region/AP by the corresponding number of notified cases yielded an estimate for the total numbers of Omicron cases in Italy in the three dates of the surveys. These were fitted with an exponential curve having growth rate *r*; we then estimated the doubling time of Omicron as *T=log(2)/r* and the net reproduction number as *R=1+r*GT*, where GT represents the average generation time. In the absence of estimates of GT for Omicron, we considered values between 4 and 8 days, covering a range of estimates for previous lineages [2–6].

We estimated an average daily exponential growth rate *r* for new Omicron infections of 0.22-0.26 days^−1^, corresponding to an average doubling time of 2.7–3.3 days and reproduction numbers in the range 1.86–2.04 and 2.73–3.08, when a generation time of 4 and 8 days was assumed respectively (Table 2). We note that, despite the decrease in Delta prevalence, the total number of estimated Delta cases also increased in the observation period from 9302-9484 on December 6, 2021, to 11018-14590 on January 3, 2022 (see Appendix).

The upsurge of SARS-CoV-2 transmission in Italy due to the Omicron spread was additionally evaluated by estimating daily SARS-CoV-2 net reproduction numbers using data collected by the Italian Integrated Surveillance System. Separate estimates were obtained from the time series of symptomatic cases by date of onset (R_sym_), and from the time series of patients admitted to hospital (R_hos_) and to intensive care units (R_ICU_) by date of admission. Methodological details are described in Appendix. R_sym_ increased from 1.15 (95% Credible Intervals, CrI: 1.15-1.15) on December 6, 2021, to a peak of 1.83 (95%CrI:1.82-1.84) on December 28 (Figure 2). R_hos_ increased from 1.14 (95%CI: 1.11-1.18) on December 6, 2021, to a peak of 1.33 (95%CrI: 1.30-1.36) on December 29, 2021. R_icu_ did not show a clear temporal trend, with mean estimates ranging between 1.09 and 1.21.

**Figure 2.**
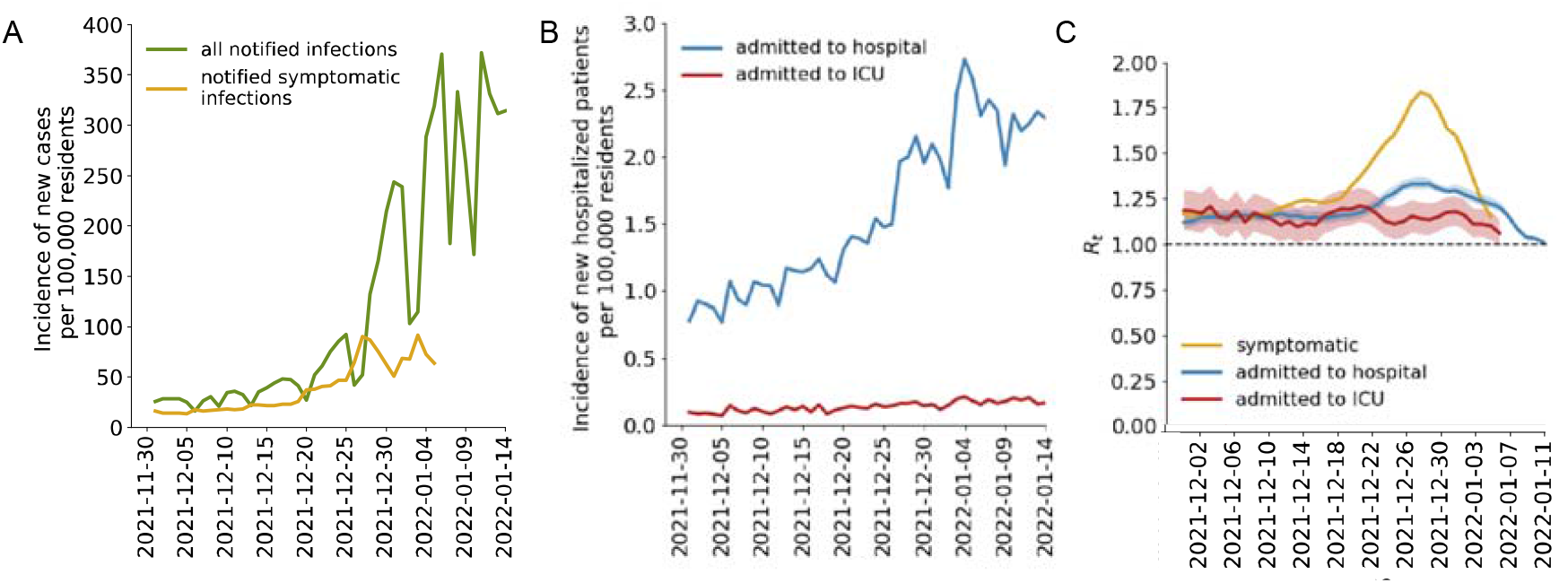
The impact of Omicron spread on SARS-CoV-2 circulation. A) Time series of total number of notified and symptomatic cases by date of notification and symptom onset, respectively. B) Time series of total number of cases admitted to hospital and to intensive care unit (ICU), by date of admission. C) Net reproduction numbers R_t_ for symptomatic cases, cases admitted to hospital and to ICU, respectively in yellow, blue and red. The solid line represents the mean of the estimated posterior distribution, and the shaded areas delimit 95% credible intervals.

## Discussion

Omicron was first identified in Italy in the second half of November 2021 [7]. Three genomic surveys were successively conducted to evaluate the progressive spread of the variant in the country. The presented results shows that Omicron became dominant across the Italian territory in less than one month, significantly increasing SARS-CoV-2 transmission.

Our estimates for the doubling time (2.7-3.3 days) are in agreement with those obtained in other countries [8,9]. The reproduction number associated to the Omicron variant under the transmissibility conditions existing in Italy in December may be in the range of 1.8-3.1. This estimate is compatible with values of R_sym_ around 1.8 observed on December 28, 2021, when the replacement of Delta by Omicron was not complete and before control measures (decided on December 23 and 29) and behavior change had a chance to significantly affect transmission. Notably, changes in contact patterns due to the Christmas holidays may have had an effect that is hard to quantify. The lower increase of reproduction numbers associated to hospital admissions and the stable value of those from ICU admissions point to a reduced clinical severity of Omicron compared to Delta, as previously suggested [10,11]. Of note, we estimate the total number of Delta cases to have increased over the study period, suggesting that a considerable fraction of hospital patients may still be due to Delta. Continued monitoring of Delta prevalence is therefore recommended.

The three daily prevalence surveys were based on relatively small sample sizes and, despite the randomization of sampling for genomic sequencing, biases due to the presence of large clusters of cases in a region cannot be excluded. Moreover, estimates of the net reproduction numbers are based on the strong assumption that the Omicron generation time is comparable to that of pre-circulating strains [2]. The selective advantage of Omicron over Delta may be explained by increased transmissibility, partial immune escape, or a combination of both [12]. Early evidence from statistical analysis of reinfections [13,14] and breakthrough infections [8,15] suggests a significant capability of escape from natural and vaccine-acquired immunity. However, further efforts are needed to quantify the relative transmissibility and clinical severity of Omicron and its immune escape ability.

## Data Availability

All data produced in the present study are available upon reasonable request to the authors

## Ethical statement

Ethical approval for sequencing SARS-CoV-2 genomes on clinical samples was obtained by ISS (ref. PRE BIO CE n.26259, July 29, 2020).

## Acknowledgements

SM, PPe, GG, and FR acknowledge funding from EU grant 874850 MOOD (catalogued as MOOD 000). The funders had no role in study design, data collection and analysis, decision to publish, or preparation of the manuscript.

## Authors’ contributions

PS, ATP, SB, GR, PPe, SM conceived the study. PPo, FT, MM wrote the first draft of the manuscript. AM, FT, PPo, MM, GG analyzed the data. AM, MSS, FM, DP, AB, FR, LA, ADM and the members of the Italian Integrated Surveillance of COVID-19 Study Group collected the epidemiological data. The members of the Genomic SARS-CoV-2 National Surveillance Working Group collected the samples and sequenced the RNA for the molecular typing. All authors revised critically the study and the results.

