## Appendix for "Tracking the progressive spread of the SARS-CoV-2 Omicron variant in Italy, December 2021 - January 2022"

**Genomic SARS-CoV-2 national surveillance working group:** Alessandra Lo Presti, Dept. Infectious Diseases, Istituto Superiore di Sanità, Rome; Stefano Morabito, Gabriele Vaccari, Ilaria Di Bartolo, Arnold Knijn, Luca De Sabato, Dept. Food safety, nutrition and veterinary public health, Istituto Superiore di Sanità, Rome. Liborio Stuppia, Laboratorio di Genetica Molecolare, Center for Advanced Studies and Technology (CAST), Università degli Studi "G. d'Annunzio", Chieti; Giovanni Savini, Istituto Zooprofilattico Sperimentale dell'Abruzzo e del Molise "Giuseppe Caporale", Teramo; Antonio Picerno, Teresa Lopizzo, UOC Analisi Chimico Cliniche e Microbiologiche, AOR "San Carlo", Potenza; Domenico Dell'Edera, UOSD Laboratorio di Genetica Medica, P.O. "Madonna delle Grazie", Matera; Pasquale Minchella, SOC Microbiologia e Virologia, AO "Pugliese-Ciaccio", Catanzaro; Francesca Greco, UOC Microbiologia e Virologia, PO "Annunziata", Cosenza; Giuseppe Viglietto, Laboratorio di Genomica Funzionale e Patologia Molecolare, Università degli Studi "Magna Graecia", Catanzaro; Maria Teresa Fiorillo, Azienda Sanitaria Provinciale di Reggio Calabria, Reggio Calabria; Luigi Atripaldi, AORN "Azienda Sanitaria dei Colli", Napoli; Antonio Limone, Istituto Zooprofilattico Sperimentale del Mezzogiorno, Napoli; Davide Cacchiarelli, Telethon Institute of Genetics and Medicine, Pozzuoli; Pierlanfranco D'Agaro, SC UCO Igiene e Sanità Pubblica, Azienda Sanitaria Universitaria Giuliano-Isontina (ASUGI), Trieste; Danilo Licastro, Laboratorio Genomica ed Epigenomica, Area Science Park, Basovizza, Trieste; Stefano Pongolini, Unità di Analisi del Rischio ed Epidemiologia Genomica, Istituto Zooprofilattico Sperimentale della Lombardia e dell'Emilia-Romagna, Parma; Tiziana Lazzarotto, Giada Rossini, Laboratori CRREM, UO

Microbiologia. AOU “Policlinico di S’Orsola” (IRCCS), Bologna; Vittorio Sambri, Dipartimento di Medicina Specialistica Diagnostica e Sperimentale (DIMES), Università di Bologna, Bologna, & UOC Microbiologia, AUSL della Romagna, Cesena; Giorgio Dirani, Silvia Zannoli, UOC Microbiologia, AUSL della Romagna, Cesena; Paola Affanni, Maria Eugenia Colucci, Laboratorio di Igiene e Sanità Pubblica, Dipartimento di Medicina e Chirurgia, Università degli Studi di Parma, Parma; Maria Rosaria Capobianchi, Laboratorio di Virologia, Istituto Nazionale Malattie Infettive IRCCS “L. Spallanzani”, Roma; Patricia Alba, Department of General Diagnostics, Department of Virology, Istituto Zooprofilattico Sperimentale del Lazio e della Toscana (IZSLT), Roma; Alice Massacci, IRCSS “Regina Elena” National Cancer Institute, Roma; Carlo Federico Perno, Unità Complessa di Microbiologia ed Immunologia Diagnostica, Ospedale Pediatrico “Bambino Gesù”, Roma; Maurizio Sanguinetti, Istituto di Microbiologia e Virologia, Fondazione Policlinico Universitario “A. Gemelli” IRCCS, Roma; Bianca Bruzzone, Laboratorio di Riferimento Regionale per la diagnosi molecolare di SARS-CoV-2, U.O. Igiene, Ospedale Policlinico “San Martino” IRCCS, Università degli Studi di Genova, Genova, & Laboratorio di Riferimento Regionale per le Emergenze di Sanità Pubblica (LaRESP), Liguria; Giancarlo Icardi, Flavia Lillo, Andrea Orsi, Laboratorio di Riferimento Regionale per le Emergenze di Sanità Pubblica (LaRESP), Liguria; Elena Pariani, Dipartimento di Scienze Biomediche per la Salute, Università degli Studi di Milano, Milano; Fausto Baldanti, Unità Virologia Molecolare, Fondazione IRCCS Policlinico “San Matteo”, Pavia, & Università di Pavia, Pavia; Maria Rita Gismondo, Valeria Micheli, U.O.C Microbiologia Clinica, Virologia e diagnostica delle Bioemergenze, ASST “Fatebenefratelli-Sacco”, Milano; Fabrizio Maggi, SC Laboratorio Microbiologia, ASST “Sette Laghi”, Varese; Arnaldo Caruso, Laboratorio di Microbiologia e Virologia, ASST “Spedali Civili di Brescia”, Brescia; Ferruccio Ceriotti, Fondazione IRCCS “Ca' Granda” Ospedale Maggiore Policlinico di Milano, Milano; Maria Beatrice Boniotti, Ilaria Barbieri, Istituto Zooprofilattico Sperimentale della Lombardia e dell'Emilia Romagna, Brescia; Alice Nava, ASST Grande Ospedale Metropolitano Niguarda, Milano; Erminio Torresani, IRCCS Istituto Auxologico Italiano, Milano; Fabiana Cro, SYNLAB ITALIA, Brescia; Enzo Boeri, Laboratorio di microbiologia, Dipartimento di Medicina di Laboratorio, Ospedale “San Raffaele”, Milano; Marina Noris, Istituto di Ricerche Farmacologiche “Mario Negri” IRCCS, Milano; Giulia Bassanini, Laboratorio SMeL, PTP Science Park S.c.a.r.l., Lodi; Claudio Farina, Marco Arosio, Laboratorio di Microbiologia e Virologia, ASST “Papa Giovanni XXIII”, Bergamo; Sergio Malandrin, Annalisa Cavallero, Laboratorio di Microbiologia e Virologia, ASST Monza, Monza; Patrizia Bagnarelli, Stefano Menzo, SOD Virologia, AOU “Ospedali Riuniti”, Ancona; Silvio Garofalo, Massimiliano Scutellà, UOC Laboratorio Analisi, POA “Cardarelli”, Campobasso; Elisabetta Pagani, Laboratorio Aziendale di Microbiologia e Virologia, Azienda Sanitaria dell’Alto Adige, Bolzano; Lucia Collini, Microbiologia e Virologia, Presidio Ospedaliero “Santa Chiara”, Trento; Valeria Ghisetti, Centro di Riferimento Regionale per validazione e controllo di qualità SARS-CoV-2, Ospedale Amedeo di Savoia, Torino; Silvia Brossa, IRCCS Fondazione del Piemonte per l'Oncologia, Candiolo; Giuseppe Ru, Elena Bozzetta, Istituto Zooprofilattico Sperimentale del Piemonte, Liguria e Valle d’Aosta, Torino; Maria Chironna, Laboratorio di Epidemiologia Molecolare e Sanità Pubblica, AOUC Policlinico di Bari, Bari; Antonio Parisi, Istituto Zooprofilattico Sperimentale della Puglia e della Basilicata, Putignano; Salvatore Rubino, Caterina Serra, S.C. Microbiologia e Virologia, Laboratorio Virologia, AOU di Sassari, Sassari; Gabriele Ibba, AMES Centro Polidiagnostico Strumentale S.r.l., AOU di Sassari, Sassari; Giovanna Piras, UOC Ematologia, P.O. "San Francesco", Azienda

Tutela Salute, ASSL Nuoro, Nuoro; Giuseppe Mameli, Laboratorio di Patologia Clinica, P.O. "San Francesco", Azienda Tutela Salute, ASSL Nuoro; Ferdinando Coghe, Laboratorio Generale (HUB) di analisi chimico cliniche e microbiologia, PO "Duilio Casula", AOU di Cagliari, Cagliari; Francesco Vitale, Fabio Tramuto, Laboratorio di Riferimento Regionale per la Sorveglianza Epidemiologica e Virologica del PROMISE - AOUP "Giaccone", Palermo; Guido Scalia, Concetta Ilenia Palermo, Laboratorio di Virologia Clinica, AOUP "V. Emanuele", PO "Gaspare Rodolico", Catania; Giuseppe Mancuso, UOC Microbologia, AOU "G. Martino ", Messina; Teresa Pollicino, Laboratorio di Diagnostica Molecolare dell'Unità Gestione Centralizzata Laboratori, Messina; Francesca Di Gaudio, Centro Regionale per la Qualità (CRQ), Palermo; Stefano Vullo, Stefano Reale, Istituto Zooprofilattico Sperimentale della Sicilia, Palermo; Maria Grazia Cusi, UOC Microbiologia e Virologia, Azienda Ospedaliera Universitaria Senese, & Dipartimento di Biotecnologie Mediche, Università degli Studi di Siena, Siena; Gian Maria Rossolini, SOD Microbiologia e Virologia, AOU "Careggi", Firenze; Mauro Pistello, UOC Virologia, AOU Pisana, Pisa; Antonella Mencacci, Barbara Camilloni, S.C. Microbiologia, Dipartimento di Medicina e Chirurgia, Università di Perugia, Perugia; Silvano Severini, Istituto Zooprofilattico Sperimentale dell'Umbria e delle Marche, Perugia; Massimo Di Benedetto, Laboratorio Analisi Cliniche, Ospedale "Parini", Aosta; Calogero Terregino, Isabella Monne, Istituto Zooprofilattico Sperimentale delle Venezie, Legnaro; Valeria Biscaro, UOC Microbiologia-Virologia, AULSS2 La Marca, PO Treviso, Treviso.

**Italian Integrated Surveillance of COVID-19 Study Group:** Del Manso Martina, Spuri Matteo, Sacco Chiara, Fabiani Massimo, Bressi Marco, Mateo-Urdiales Alberto, Vescio Maria Fenicia, Istituto Superiore di Sanità, Rome, Italy.

### Sample size estimation for genomic surveillance

Genomic surveillance in Italy is coordinated by the Italian National Institute of Health, in collaboration with the Ministry of Health and the laboratories of Regions/Autonomous Provinces (AP). Within the context of rapid detection and surveillance of emerging variants of concern (VOC), ad-hoc surveys are carried out in Italy. From the emergence of the VOC Omicron, three genomic surveys were conducted on December 6, 2021, December 20, 2021, and January 3, 2022, to estimate its prevalence. As of December 6, Delta was the predominant SARS-CoV-2 lineage in Italy [1].

The surveys involved all the 19 Regions and 2 Autonomous Provinces (AP) of Italy. Random samples of SARS-CoV-2 positive cases were analyzed by 120 laboratories distributed across the national territory. Samples were selected from both passive surveillance and contact tracing and included symptomatic, pre-symptomatic and asymptomatic cases. The collected samples were sequenced according to the local laboratory policy by either of the following techniques: i) sequencing the entire S-gene by Sanger technology, ii) sequencing part of the S-gene with the identification of all mutations/deletions associated with the three variants, or iii) sequencing the whole genome by Next Generation Sequencing.

Samples were distributed across 4 macro-areas, defined according to the Eurostat NUTS1 classification: North-East, North-West, Center, and South/Islands. The sample size was calculated to have the statistical power to detect a prevalence of 5%, with precision 2%, within each macro-area, based on the following equation

$$n \geq \frac{\mathrm{N} z_{\alpha/2}^{2}\, \mathrm{p}(1 - \mathrm{p})}{\varepsilon^{2}(\mathrm{N} - 1) + z_{\alpha/2}^{2} \mathrm{p}(1 - \mathrm{p})}$$

where N is the total number of cases notified on the day preceding each of the three surveys [2], $p$ is the target prevalence (5%), $\varepsilon$ is the desired precision (2%), $\alpha$ is the significance level set at 5%, and $z_{\alpha/2}^{2}$ the corresponding z-score (1.96).

### Estimation of Omicron prevalence and notified cases from surveys data

To estimate the Omicron prevalence and the number of notified cases, we employed two alternative approaches that are described below.

*Monte Carlo Markov Chain approach*

We fitted the Omicron prevalence in the three surveys by means of a Markov Chain Monte Carlo (MCMC) approach applied to the binomial likelihood of observing the identified number of omicron infections among the sequenced genomes. The MCMC approach was applied considering two different subdivisions of the Italian territory. In the first one, we consider regions as the observational unit; in the second one, the 4 macro-areas defined in the previous section. We fitted 10000 iterations of MCMC separately for each unit (either region or macro-area) and for each survey. We then approximated the local number of Omicron cases by multiplying the obtained prevalence with the corresponding number of cases notified at the date of sample collection. Secondary outcomes are *i)* the distributions of the number of notified Omicron cases in each unit and in the whole Italy (obtained by addition of local cases) and *ii)* the distributions of Omicron prevalence in the whole Italy, obtained by dividing the estimated number of Omicron cases by the total number of cases notified on 6 December 2021, 20 December 2021, and 3 January 2022. The same approach was used to approximate the number of notified Delta cases over the three survey days. Results are reported in the main text and in Table S2.

*Regression modelling approach*

We fitted a generalized linear mixed model to estimate Omicron prevalence when the three surveys were conducted. We applied a Bayesian approach for the estimation of model parameters. The model is applied considering two different subdivisions of the Italian territory. In the first one, we consider regions as the observational unit; in the second one, we aggregated regional data in the 4 macro-areas defined in the previous section. In both cases, we assumed a binomial distribution for the dependent variable, represented by the total number of omicron sequences out of the total number of sequences analyzed in each unit (either region or macro-area). We considered as independent variable the date when surveys were carried out. Dates were expressed as the number of days elapsed from the first survey: 6 December 2021 corresponds to day 0; 20 December 2021 to day 14; 3 January 2022 to day 28. For computational convenience, we rescaled the day variable by dividing it by its maximum value. We considered a random intercept and a random slope for each unit (either region or macro-area) to allow for heterogeneities among units both in the detected initial prevalence and in the growth rate of Omicron prevalence. The adopted model equation was:

$$y_{i,s} \sim Bin(p_{i,s}, S_{i,s})$$
$$logit(p_{i,s}) = \alpha + a_i + (\beta + b_i)\, day_s$$
$$\alpha \sim Norm(-5, 0.1)$$
$$\beta \sim Norm(5, 1000)$$
$$a_i \sim half - Cauchy(25)$$
$$a_i \sim half - Cauchy(25)$$

where the subscript *i* and *s* identify the geographical unit and the survey considered, respectively; $\alpha$ is the intercept, representing the average prevalence among units, and is assumed to follow an informative Normal prior of parameters mean -5 and variance 0.1; $\beta$ is the slope of the regression model, representing the average rate of change in prevalence by day among units, and it is assumed to follow a non-informative Normal prior distribution of parameters mean 5 and variance 1000; $a_i$ and $b_i$ are the random effect coefficients for the intercept and the slope, respectively, and are assumed to follow a half-Cauchy prior of parameter 25; $day_s$ represents the number of days from the first survey to survey *s* (which is the same for all units *i*). $S_{i,s}$ represents the total number of sequences analyzed in unit *i* during survey *s*; $y_{i,s}$ is the number of Omicron sequences confirmed in unit *i* during survey *s* and $p_{i,s}$ is the estimated Omicron prevalence in unit *i* during survey *s*. Three chains were used in the MCMC process with a burn-in of 100000 iteration and a thinning rate of 50 resulting in 3000 iterations used to estimate each posterior distribution.

The posterior distributions of the number of notified Omicron cases in each unit and in the entire Italy (by addition) on 6 December 2021, 20 December 2021, and 3 January 2022 were obtained by multiplying the estimated posterior distribution of Omicron prevalence with the corresponding number of cases notified at the date of sample collection. The same approach was used to approximate the number of notified Delta cases over the three survey days. Results are reported in the main text and in Table S2.

### Estimation of Omicron exponential growth rate, doubling time and net reproduction number (*R*) from surveys data

To estimate the growth rate, the doubling time and R associated with Omicron infections and its potential impact on SARS-CoV-2 transmissibility, we applied a linear regression model to a sample (of size 1000) pooled from the posterior distribution of Omicron cases at national level notified at the date of sample collection. The posterior distribution of Omicron cases was estimated through the approaches described in the previous paragraphs. For each triplet of log-transformed cases we applied the following linear regression model:

$$logC_s \sim Norm(\mu_s, \sigma_s^2)$$
$$\mu_s = \eta \ + \psi \ day_s$$
$$\eta \sim Norm(logC_1, 1)$$
$$\psi \sim Norm((logC_3 - logC_1)/day_3 \ , 0.1)$$
$$\sigma_s \sim Gamma(1.5,2)$$

where $logC_s$ is the log-transformed number of notified Omicron cases in Italy on the day of survey *s*, which is assumed to follow a normal distribution of mean $\mu_s$ and standard deviation $\sigma_s$. We assumed a gamma distribution of shape 1.5 and rate 2 for $\sigma_s$. $\eta$ and $\psi$ are the intercept and the slope in the regression equation, respectively. The slope $\psi$ should be interpreted as the growth rate *r* of the epidemic. We assumed for $\eta$ an informative normal prior distribution of mean equal to the log-transformed number of notified cases on the day of the first survey and standard deviation equal to 1. We assumed for $\psi$ an informative normal prior distribution of mean equal to the slope of the line intersecting the log-transformed number of notified cases during the day of the first and third survey and standard deviation equal to 0.1. Three chains were used in the MCMC process with a burn-in of 150000 iteration and a thinning rate of 10 resulting in 3000 iterations used to estimate each posterior distribution.
Finally, we computed the posterior distribution of *r* by sampling 1000 values from each one of the posterior distributions of $\psi$ obtained from the pooled 1000 triplets of cases.

The estimated distribution of the exponential growth rate *r* was used to obtain the distribution of the doubling time *T* and of the net reproduction number (*R*) expected after the Omicron expansion in Italy through the following formulas

$$T = \log(2)\ /r$$
$$R = 1 + r * GT$$

Where GT represents the Omicron generation time (i.e., the average time elapsing between the infection of primary and secondary cases). In the absence of robust estimates for the generation time of Omicron, we considered values of 4, 6 and 8 days.

### Estimation of the SARS-CoV-2 net reproduction numbers (*Rt*) from surveillance of cases

To evaluate the progressive impact of the Omicron expansion on the overall SARS-CoV-2 circulation, we estimated the time varying net reproduction number *Rt* associated to cases reported to the National Integrated Surveillance System. The approach adopted to estimate *Rt* is the same used during the official weekly monitoring of SARS-CoV-2 transmission in Italy. Methodological details can be found in [3]. The net reproduction number quantifies the transmission potential at a given time *t* and is used to monitoring exogenous and endogenous changes in the infection transmission, including those related to performed control measures, changes in the susceptibility and in human behavioral responses to the infection risks.

**Software**

All the analysis were performed using the statistical software *R* (version 4.1.0) [4] and *JAGS* (version 4.3.0) [5], and the related packages *R2Jags* [6], *here* [7] and *tidyverse* [8].

**Table S1. National level estimates from surveys data for the Omicron and Delta number of cases**

| | | GLMM | | MCMC | |
|---|---|---|---|---|---|
| | | **By region** | **By macro area** | **By region** | **By macro area** |
| **Omicron cases** | **December 6th, 2021** | 99 (72 - 130) | 104 (76 - 134) | 135 (82 - 201) | 45 (19 - 83) |
| | **December 20th, 2021** | 3039 (2780 - 3307) | 3103 (2842 - 3369) | 3489 (3193 - 3809) | 3481 (3196 - 3775) |
| | **January 3rd, 2022** | 55920 (54816 - 57034) | 55302 (54226 - 56342) | 54612 (53462 - 55715) | 54501 (53469 - 55524) |
| **Delta cases** | **December 6th, 2021** | 9404 (9373 - 9431) | 9399 (9369 - 9427) | 9368 (9302 - 9421) | 9458 (9420 - 9484) |
| | **December 20th, 2021** | 13174 (12906 - 13433) | 13110 (12844 - 13371) | 12724 (12404 - 13020) | 12732 (12438 - 13017) |
| | **January 3rd, 2022** | 12132 (11018 - 13236) | 12750 (11710 - 13826) | 13440 (12337 - 14590) | 13551 (12528 - 14583) |
